# Development and Validation of a Multimodal AI-Based Model for Predicting Post-Prostatectomy Treatment Outcomes from Baseline Biparametric Prostate MRI

**DOI:** 10.64898/2026.03.19.26348716

**Authors:** Benjamin D. Simon, Esra Akcicek, Stephanie A. Harmon, Lei Clifton, Anshul Thakur, Sandeep Gurram, David Clifton, Bradford J. Wood, Ali Devrim Karaosmanoglu, Peter L. Choyke, Deniz Akata, Peter A. Pinto, Baris Türkbey

## Abstract

Prostate cancer (PCa) is the second most common cancer and cause of cancer death in American men. Existing risk prediction methods have limited accuracy and reproducibility, resulting in difficulty in predicting disease severity. We demonstrate the development and external validation of an automated multimodal artificial intelligence algorithm using biparametric MRI (bpMRI) and clinical covariates for predicting biochemical recurrence (BCR) after radical prostatectomy (RP) in PCa patients. Development cohort included 80% of patients from center 1 (*n* = 240) who underwent prostate MRI prior to RP between January 2008 and December 2018 with a minimum of two years of follow-up after RP. Test cohort included the remaining 20% of center 1 patients (*n* = 71), and the external validation cohort from center 2 (*n* = 168). Center 2 patients included those who underwent prostate MRI and RP between January 2015 and December 2024 with a minimum of two years of follow-up. Clinical comparisons were CAPRA-S (center 1) and ISUP grade group from post-RP biopsy (center 2). Models developed were a clinical model (M0), an automated clinical model (M1), a radiomics model (M2), and a multimodal model (M3). Clinical variables (M0) included PSA, age, primary Gleason, and ISUP grade group. Automated clinical variables (M1 and M3) included PSA and age. Radiomics features (M2 and M3) were extracted from bpMRI using a lesion detection algorithm. Accuracy, sensitivity, specificity, and AUC were calculated, and log-rank tests compared BCR-free survival to assess the models’ ability to discriminate relative to clinical standards. Intermediate-risk groups were also assessed. The multimodal model (M3) had the highest AUC across test sets (combined: 0.71; center 1: 0.70; center 2: 0.75) and was the only model to significantly differentiate BCR-free survival outcomes in intermediate-risk groups across both centers (*p* < 0.05). This automated multimodal model leveraging radiomics and clinical covariates can predict BCR after RP, approaches clinical gold standards, and may enhance imaging-based prognostication following further validation.

## I. INTRODUCTION

Prostate cancer (PCa) is the second leading cause of cancer death among American men. In 2025, there were projected to be 313,780 new diagnoses and 35,770 cancer deaths in America and 712,000 global deaths per year in 2040 [1, 2]. Although PCa has a reputation for being treatable due to the high rates of low-grade disease and availability of blood-test screening through PSA, it is a heterogeneous disease with multiple treatment options and continued risk of metastases and mortality [3]. High-grade and distant-stage disease are on the rise despite advances in treatment technologies. There are several common tools to stratify patients’ risk of metastasis to avoid overtreatment and undertreatment. Scores such as the NCCN risk groups, Gleason grading, CAPRA-S, and PI-RADS suffer from biopsy sampling error and inter-reader disagreement. While these metrics can differentiate outcomes on a population level, for an individual patient it is unclear how sensitive or specific these scores may be. Despite their shortcomings, these metrics still inform treatment decisions, presenting an opportunity to improve outcomes by exploring a precision-medicine approach. There is a need to introduce technology that is consistent across centers and accurately prognostic on an individual patient level in order to determine optimal treatments. With MRI being relatively new to PCa guidelines, this signal-rich imaging technology provides an opportunity for modern AI and computer vision to play a role [4].

For higher-risk PCa, treatment often comes down to the decision of surgery or radiation therapy with or without androgen deprivation therapy. With roughly equivalent long-term outcomes, this decision often depends on how patients and providers choose to weigh side effects [5]. Sometimes, factors such as extraprostatic extension (EPE) or age will shift the decision away from surgery due to higher likelihood of positive margins or risk of complications [6]. These factors, too, are subjective and may vary across physicians and hospitals [7].

One solution to the inter-reader disagreement across gold-standard risk-stratification techniques is artificial intelligence (AI), which has demonstrated potential to objectively assist a variety of medical decision-making processes across multiple modalities [8]. Although there is some exploration of AI in PCa, this mostly focuses on detection, and it is difficult to find high-quality multimodal work with sufficient external validation [3, 9]. There is a need for automated multimodal pipelines to predict outcomes for RP patients with external validation for PCa. Here, we aimed to develop and evaluate a multimodal deep learning-based AI algorithm using biparametric MRI (bpMRI) and clinical covariates for predicting biochemical recurrence (BCR) after RP in prostate cancer patients.

## II. METHODS

### A. Development Cohort: Center 1 Patient Population

This retrospective study was institutional review board approved and HIPAA compliant (ClinicalTrials.gov identifiers NCT02594202 and NCT03354416). Patients who underwent prostate MRI prior to RP between January 2008 and December 2018 (*n* = 683) were considered. Criteria for exclusion are described in detail in Fig. 1A and resulted in a final dataset of 311 patients. Biopsy methods and reader details are provided in the Supplemental Methods. Clinical covariates (age, PSA, Gleason scores, and ISUP grade group) were recorded at the time of MRI or biopsy, and follow-up for BCR was assessed using PSA values, with PSA ≥ 0.2 ng/mL defining BCR. The RP specimen was used to calculate a post-surgical CAPRA-S score for predicting BCR after surgery [10]. Data were split by pseudorandomization into *n* = 240 training and *n* = 71 test images, consistent with the split used in our previously developed open-source lesion-detection algorithm to avoid data leakage and maintain test-set independence [11]. Center 1 image-acquisition details are provided in the Supplemental Methods.

**Figure 1.**
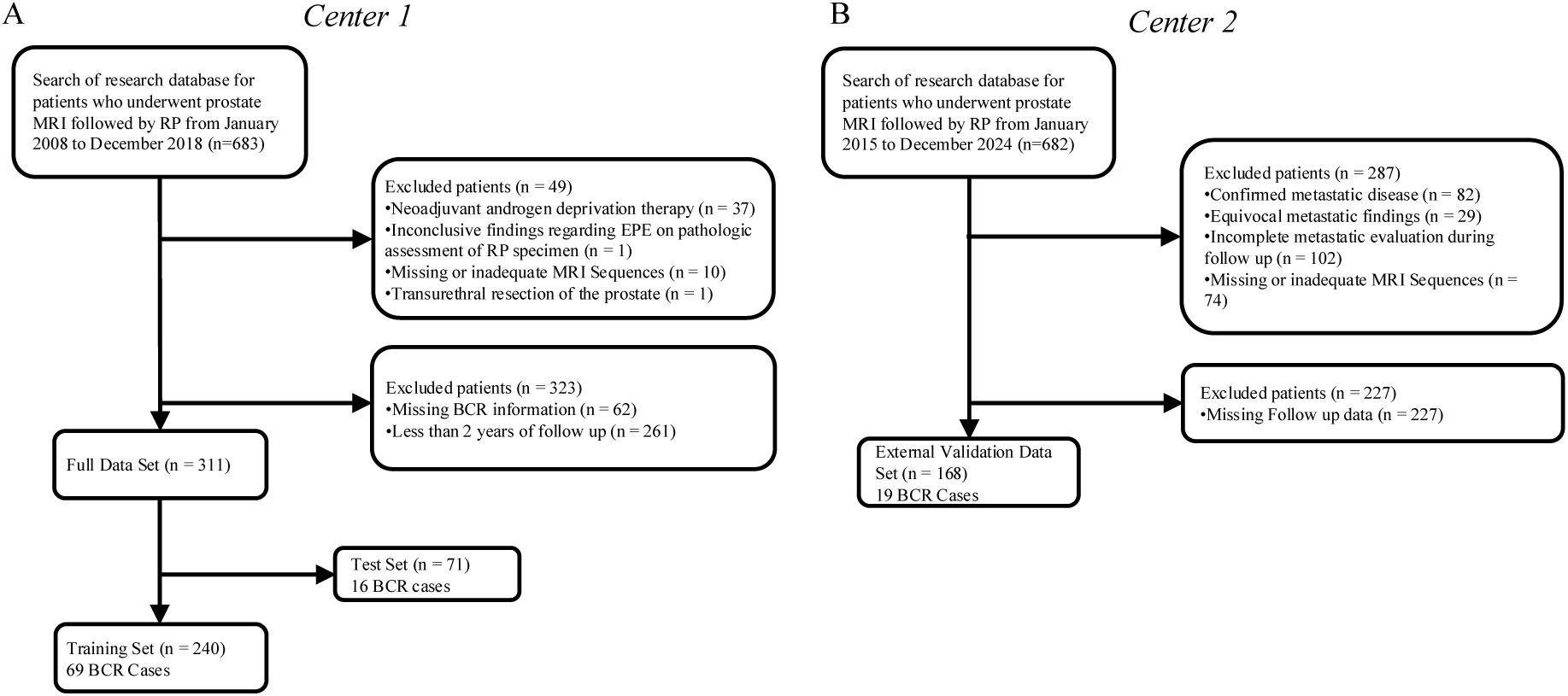
Patient population flowchart. (A) Retrospective query for center 1 patients who underwent RP during the specified interval and exclusions based on other treatments, image quality, and follow-up data. (B) Retrospective query for center 2 patients who underwent RP during a specified interval with metastatic evaluation, imaging, and follow-up data.

### B. External Validation Cohort: Center 2 Patient Population

This retrospective study was institutional review board approved (Hacettepe University IRB approval number SBA 24/174) and included de-identified data only. Patients who underwent prostate MRI prior to RP and received a diagnosis of PCa at center 2 between January 2015 and December 2024 (*n* = 682) were considered for this study. Criteria for exclusion are described in detail in Fig. 1B and resulted in a final dataset of 168 patients. Biopsy methods and reader details are in the Supplemental Methods. Clinical covariates (age, PSA, Gleason scores, and ISUP grade group) were recorded at the time of MRI or biopsy, and follow-up for BCR was based on PSA values, again using PSA ≥ 0.2 ng/mL as the definition of BCR. Center 2 image-acquisition details are provided in the Supplemental Methods.

### C. Lesion Segmentation Model and Radiomics Extraction

This model was built on a previously developed and publicly available lesion-detection and segmentation model [11]. The model leveraged T2-weighted imaging, ADC maps, and high-*b*-value diffusion-weighted imaging to produce a lesion segmentation. Subsequent work has leveraged this model for various purposes, including EPE detection, external validation, and related prostate-imaging applications [12–14]. These lesion masks were then used to measure 112 T2-weighted MRI radiomics features using PyRadiomics version 3.10.0 in Python. T2-weighted MR images were normalized, and default parameters were used for feature extraction. Features included both texture and volumetric measurements.

### D. Outcome Prediction Models

These measurements, together with clinical covariates (age and PSA), were used as inputs in XGBoost models. This pipeline is summarized in Fig. 2. Hyperparameter tuning was completed using five-fold cross-validation with AUC as the objective. Training did not consider time to BCR. Four models (M0–M3) were developed.

**Figure 2.**
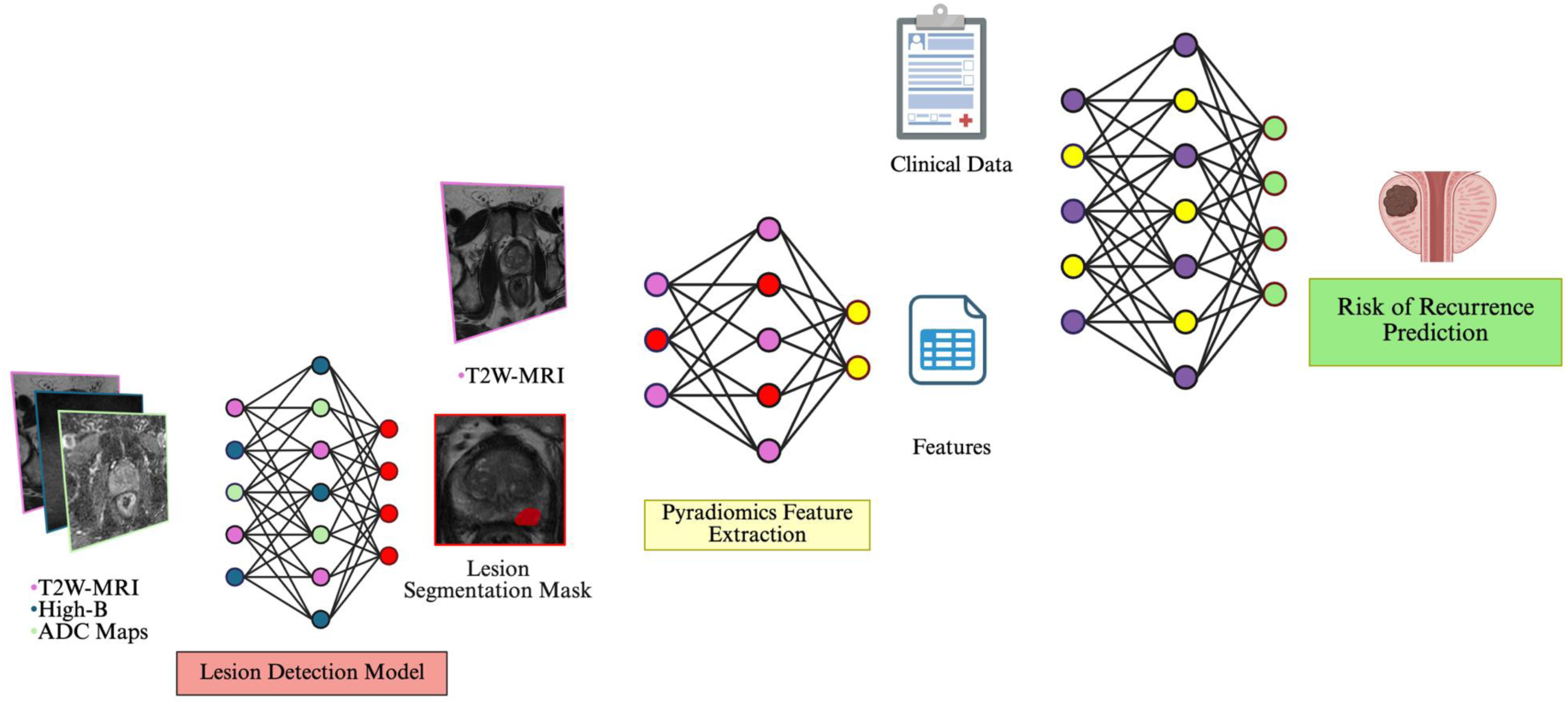
Model pipeline schematic showing the three imaging sequences used in the lesion-segmentation model, followed by T2-weighted MRI radiomics extraction with PyRadiomics and integration with clinical data to predict BCR.

M0 is a comprehensive clinical model without MRI information. Inputs included age, PSA, primary Gleason score, and ISUP grade group. This model serves as a control to evaluate M2 and M3.

M1 is a purely automated clinical model without MRI information. Inputs included only age and PSA. This model also serves as a control to evaluate M2 and M3.

M2 is a purely imaging-based model with only the 112 radiomics features associated with the deep-learning-produced lesion mask and T2-weighted MRI.

M3 is the automated multimodal model combining these imaging features with age and PSA.

The models and associated code are publicly available at https://github.com/NIH-MIP/Multimoda lRadiomicsSurgeryModel.

### E. Statistical Analysis

For each model and cohort, the area under the receiver-operating characteristic curve (AUC) was calculated. Youden’s *J* statistic in the training set was used as a cutoff for the model’s final prediction, and accuracy, sensitivity, specificity, positive predictive value, and negative predictive value are reported. Kaplan–Meier curves were generated for each cohort and for the combined cohort to evaluate prediction in the context of time to BCR. Log-rank tests were performed to compare these curves. CAPRA-S was evaluated for the development-cohort test set as a clinical gold-standard baseline, and post-RP biopsy ISUP grade group was used for the external validation cohort because margin status and regional biopsy information were unavailable. A comparison to pre-operative biopsy ISUP grade group is also included. Equivalent analyses were conducted for intermediate- and low-risk patients, for whom identifying those who will experience BCR is especially challenging.

## III. RESULTS AND DISCUSSION

### A. Patient Population: Center 1

Of the 311 patients from center 1, 85 had BCR. This constituted 29% of the training set (69/240) and 23% of the test set (16/71).

Post-surgical CAPRA-S was calculated for center 1 patients, with most receiving an intermediate CAPRA-S score of 3–5 (66% [157/240] in the training set and 72% [51/71] in the test set). Of those with BCR, a higher percentage had high-risk CAPRA-S scores of 6 or higher (54% [37/69] in the training set and 38% [6/16] in the test set).

### B. Patient Population: Center 2

Of the 168 patients in center 2, 19 had documented BCR, corresponding to 11.3% of the external validation cohort (19/168).

Post-surgical CAPRA-S could not be calculated for center 2, so the ISUP grade group at post-surgical biopsy was used as a clinical baseline. Most patients had an intermediate ISUP grade group score of 2–3 (58%, 97/168), compared with 50/168 in ISUP 1 and 21/168 in ISUP 4–5. Those with high-risk ISUP had a larger proportion of BCR events (6/21) compared with ISUP 2–3 (13/97) and low-risk ISUP 1 (2/50). Complete demographic information for each center is summarized in Table I.

**Table I.**
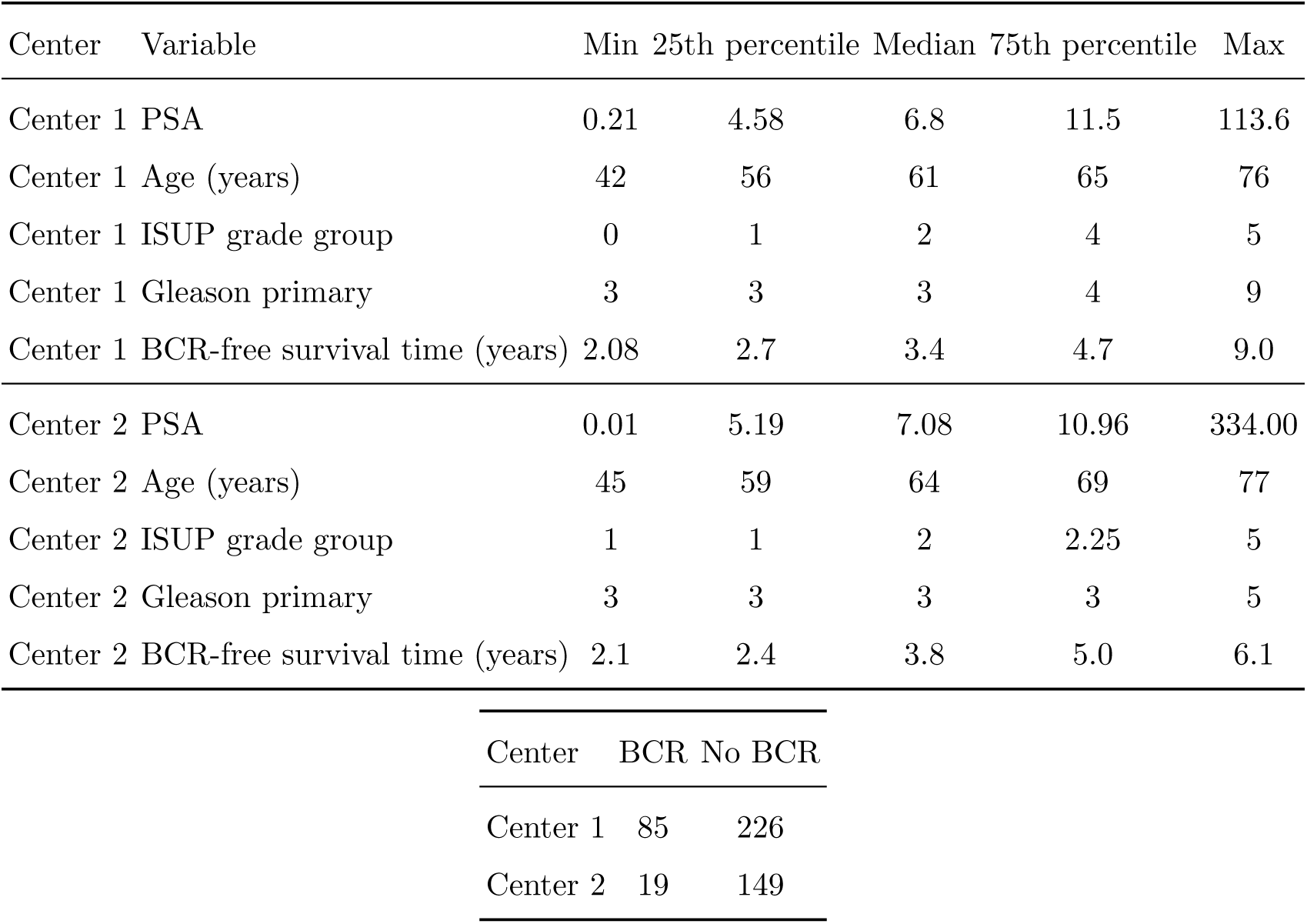
Patient characteristics. Basic clinical distribution statistics for each center and BCR event counts.

| Center | Variable | Min | 25th percentile | Median | 75th percentile | Max |
| --- | --- | --- | --- | --- | --- | --- |
| Center 1 | PSA | 0.21 | 4.58 | 6.8 | 11.5 | 113.6 |
| Center 1 | Age (years) | 42 | 56 | 61 | 65 | 76 |
| Center 1 | ISUP grade group | 0 | 1 | 2 | 4 | 5 |
| Center 1 | Gleason primary | 3 | 3 | 3 | 4 | 9 |
| Center 1 | BCR-free survival time (years) | 2.08 | 2.7 | 3.4 | 4.7 | 9.0 |
| Center 2 | PSA | 0.01 | 5.19 | 7.08 | 10.96 | 334.00 |
| Center 2 | Age (years) | 45 | 59 | 64 | 69 | 77 |
| Center 2 | ISUP grade group | 1 | 1 | 2 | 2.25 | 5 |
| Center 2 | Gleason primary | 3 | 3 | 3 | 3 | 5 |
| Center 2 | BCR-free survival time (years) | 2.1 | 2.4 | 3.8 | 5.0 | 6.1 |

| Center | BCR | No BCR |
| --- | --- | --- |
| Center 1 | 85 | 226 |
| Center 2 | 19 | 149 |

### C. Hyperparameter Tuning

Hyperparameter tuning was completed for all models based on cross-validation AUC (Table II).

**Table II.** Hyperparameter tuning selections. Distribution of hyperparameters evaluated using five-fold cross-validation of the training set and selected parameters for each model.

| Hyperparameter | Search space | M0 | M1 | M2 | M3 |
| --- | --- | --- | --- | --- | --- |
| Estimators | 100, 150, 200, 250, 300 | 250 | 200 | 100 | 200 |
| Learning rate | 0.01, 0.05, 0.1, 0.2, 0.3 | 0.05 | 0.01 | 0.3 | 0.01 |
| Maximum depth | 3, 4, 5, 6, 7, 8, 9, 10 | 5 | 5 | 5 | 5 |
| Subsample | 0.6, 0.8, 1.0 | 0.80 | 0.60 | 0.60 | 0.60 |
| Minimum child weight | 1, 3, 5 | 3 | 5 | 1 | 5 |
| Gamma | 0, 0.1, 0.2, 0.3 | 0 | 0.3 | 0.2 | 0.3 |
| Lambda | 0, 0.1, 0.5, 1, 2, 5 | 1 | 5 | 1 | 5 |
| Alpha | 0, 0.1, 0.5, 1, 2, 5 | 0.1 | 5 | 5 | 5 |
| Column subsample ratio | 0.6, 0.8, 1.0 | 0.80 | 0.60 | 0.60 | 0.60 |

### D. Model Results

For each model, performance was evaluated using AUC overall and for each cohort separately, treating BCR as a binary outcome. The AUC in the combined set of test patients from centers 1 and 2 was 0.54 for M0, 0.61 for M1, 0.65 for M2, and 0.71 for M3. Using the optimal threshold as determined by Youden’s *J* statistic in the training set, M3 achieved the highest sensitivity (94%), whereas M0 had the highest specificity (70%) and accuracy (66%). Complete binary results are shown in Table III, with AUC summaries presented in Fig. 3.

**Figure 3.**
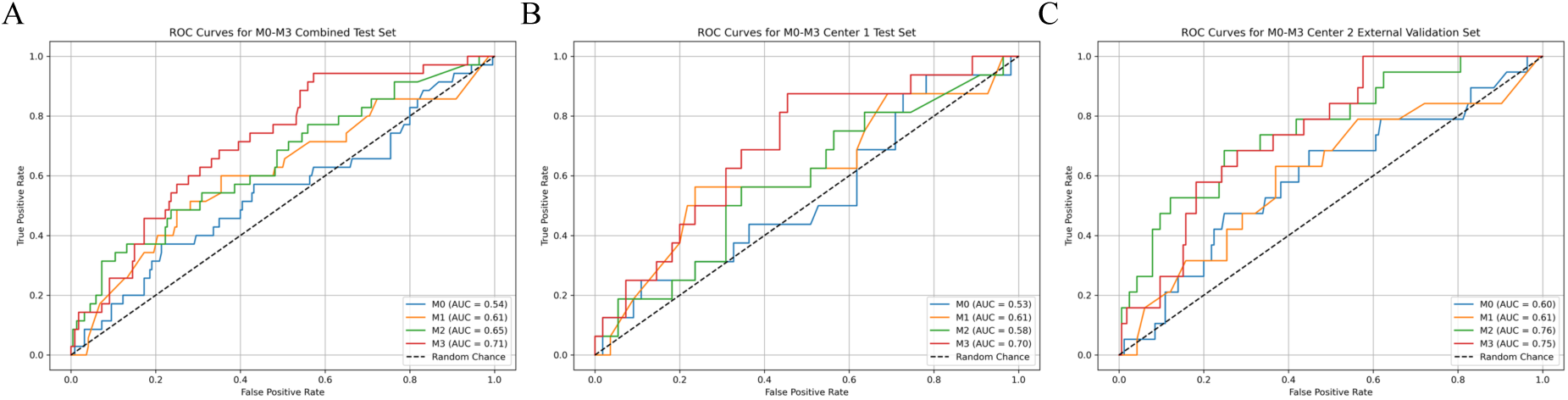
AUC results by model and cohort. (A) Combined test set. (B) Center 1 test set. (C) Center 2 external validation set.

**Table III.**
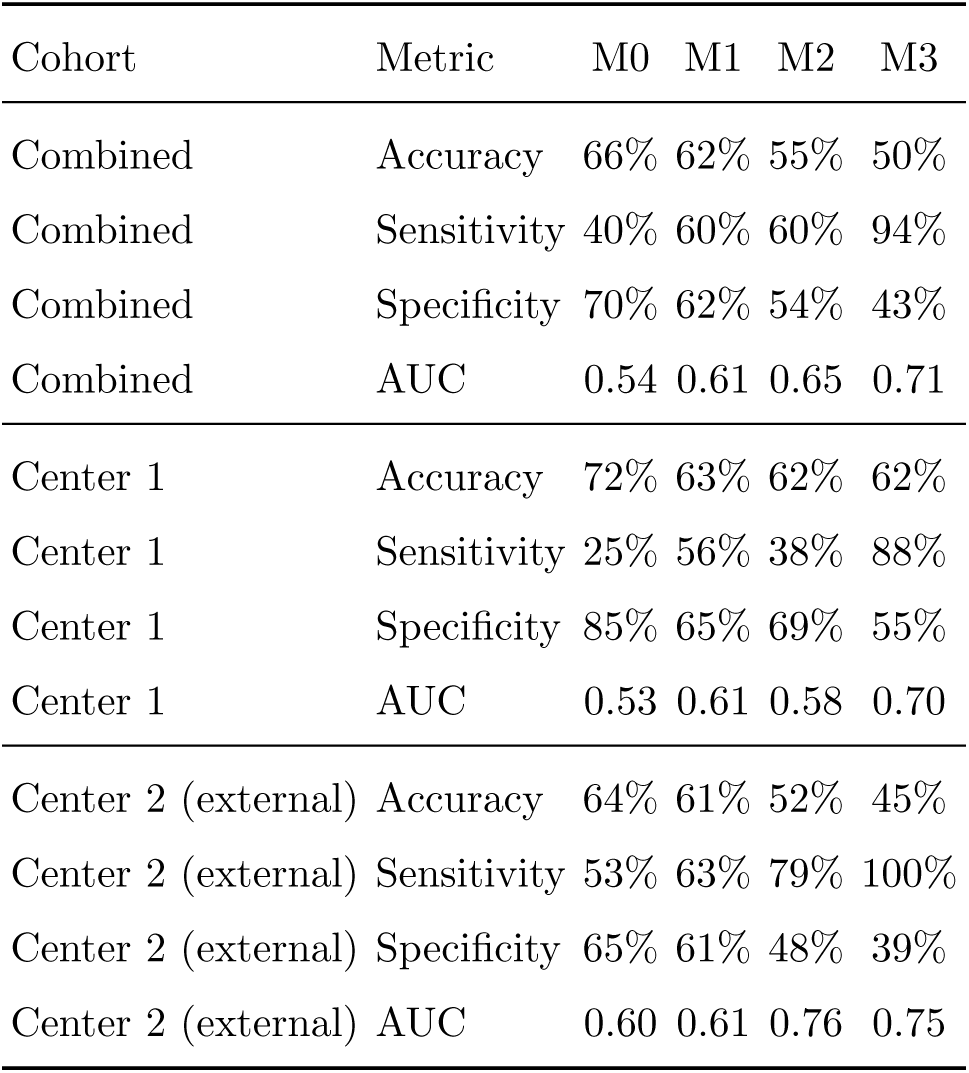
Model M0–M3 results at the optimal threshold determined from the training set.

In the center 1 cohort, the AUC was 0.53 for M0, 0.61 for M1, 0.58 for M2, and 0.70 for M3. Using the optimal threshold, M3 achieved the highest sensitivity (88%), whereas M0 had the highest specificity (85%) and accuracy (72%). In the center 2 cohort, the AUC was 0.60 for M0, 0.61 for M1, 0.76 for M2, and 0.75 for M3. Using the optimal threshold, M3 achieved the highest sensitivity (100%), while M0 had the highest specificity (65%) and accuracy (64%). The feature importances for each model are presented in Table IV.

**Table IV.** Top feature-importance weights for models M0–M3.

| M0 feature | Weight | M1 feature | Weight | M2 feature | Weight | M3 feature | Weight |
| --- | --- | --- | --- | --- | --- | --- | --- |
| PSA | 30.5% | PSA | 89.6% | GLSZM Zone Variance | 8.4% | PSA | 3.7% |
| Primary Gleason | 28.9% | Age | 10.4% | Shape Sphericity | 6.0% | Shape Least Axis Length | 2.9% |
| ISUP | 22.2% | – | – | First Order Range | 5.0% | Shape Maximum 2D Diameter Column | 2.6% |
| Age | 18.4% | – | – | GLSZM Small Area Emphasis | 3.3% | Shape Mesh Volume | 2.4% |
| – | – | – | – | GLRLM Run Length Non-Uniformity | 3.1% | GLSZM Gray Level Variance | 2.4% |

### E. Outcome Analysis

Although each model was trained on binary outcomes, time to BCR is clinically important. Accordingly, Kaplan–Meier BCR-free survival curves were generated for each model in the combined cohort, by cohort, and within intermediate-risk subgroups. A log-rank test was completed between the two prediction groups (BCR versus BCR-free) using the optimal cutoff. Statistically significant BCR-free survival differentiation was achieved for models M1 and M3 in the combined cohort. M0, M1, and M3 achieved significance in center 1. M1, M2, and M3 achieved significance in center 2 (*p* < 0.05). These survival curves are displayed in Fig. 4.

**Figure 4.**
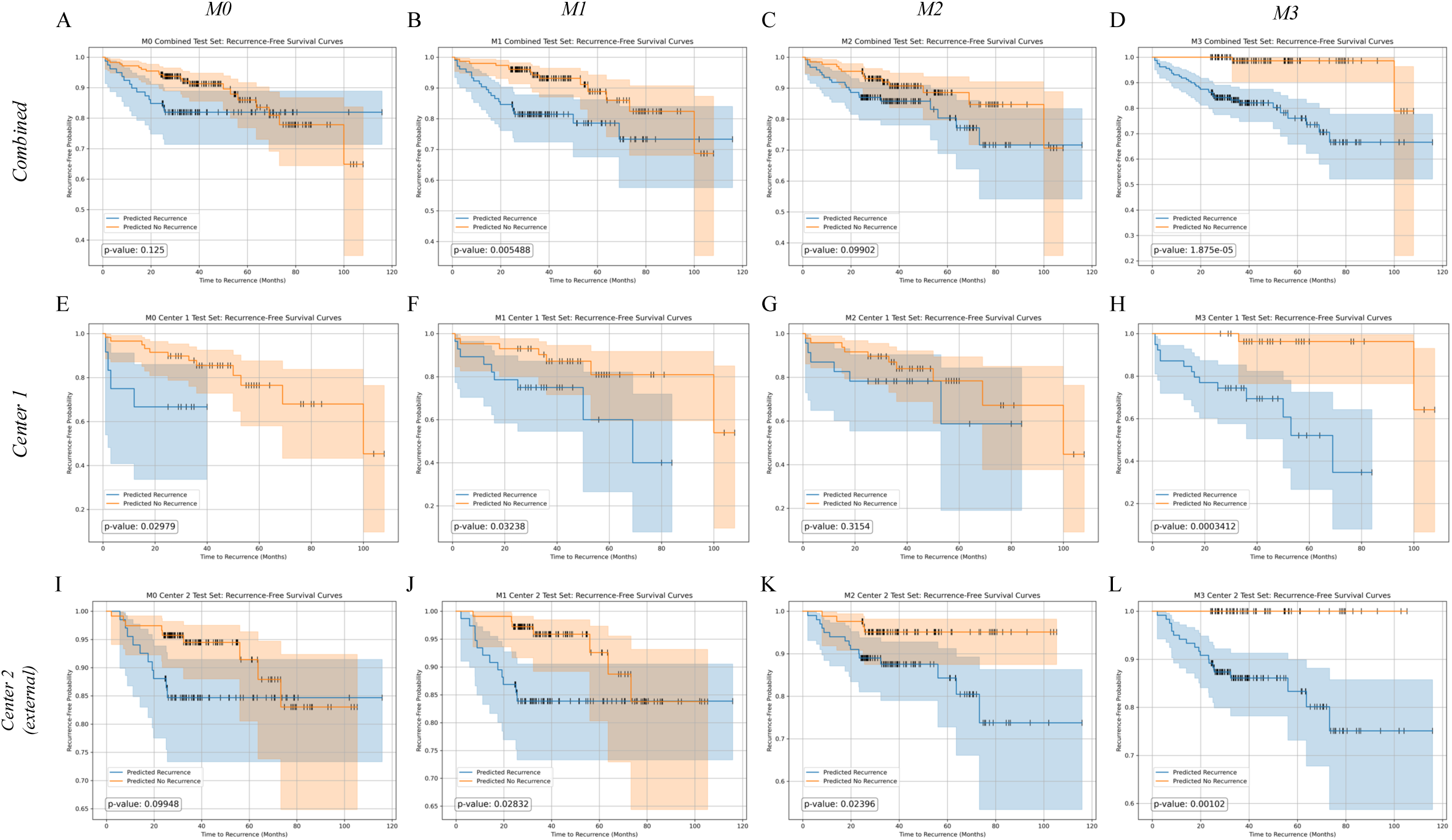
Recurrence-free survival curves by model and center with log-rank tests. Panels A–D show the combined test set; E–H show center 1; I–L show center 2.

**Figure 5.**
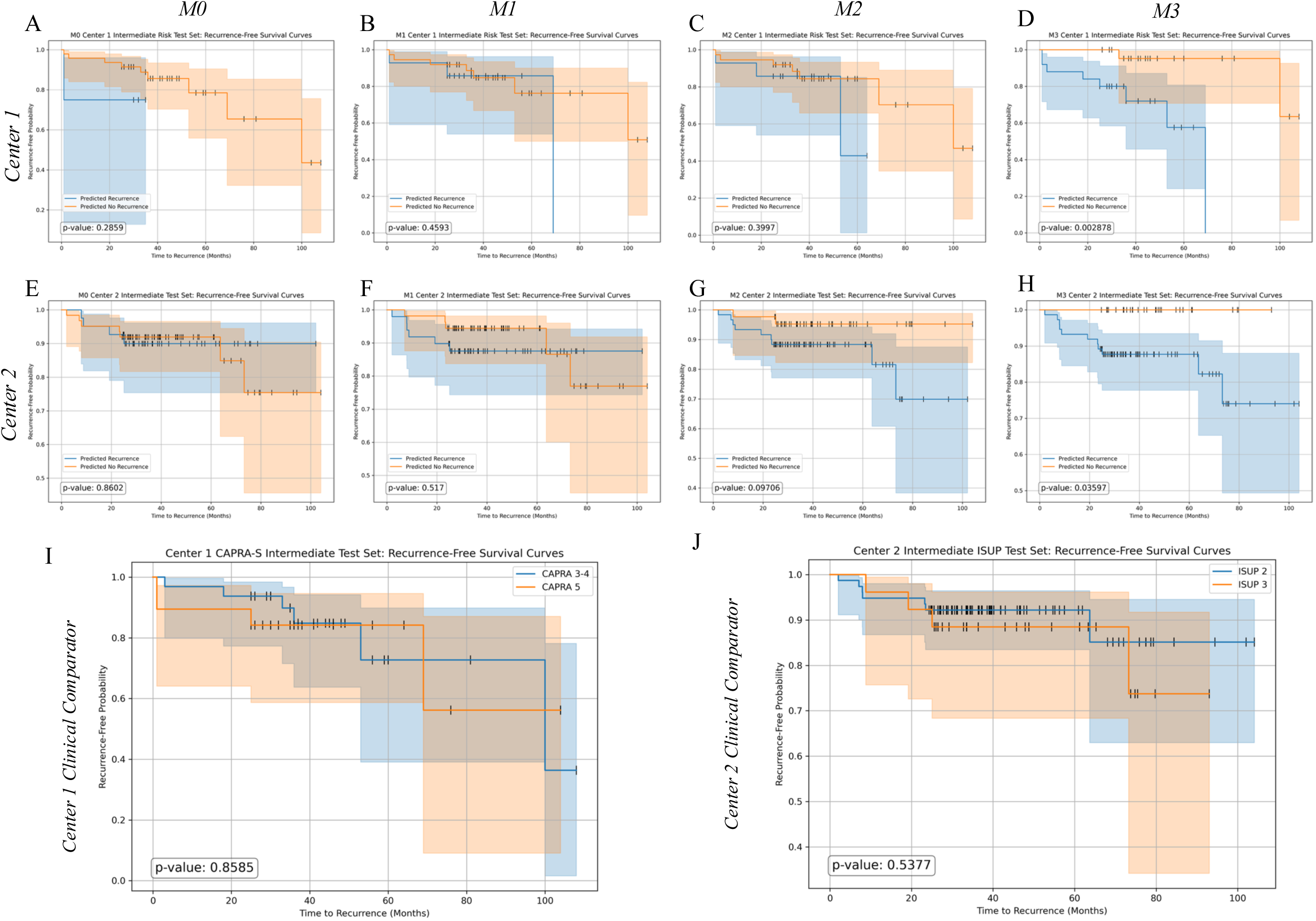
Intermediate-risk comparisons by center. Kaplan–Meier curves for intermediate-risk patients by model and center, with clinical baseline comparisons.

### F. Clinical Comparison

Because post-surgical ISUP, CAPRA-S, and related prognostic groupings generally perform relatively well in low- and high-risk individuals, it is especially important to investigate performance in intermediate-risk subgroups [4, 15]. Across all center 1 patients, CAPRA-S significantly differentiated outcomes in the small center 1 test set. For intermediate-risk center 1 patients with CAPRA-S scores 3–5, CAPRA-S itself as well as models M0, M1, and M2 did not differentiate outcomes (*p >* 0.05). However, M3 differentiated outcomes in the same cohort (*p* < 0.05).

For all center 2 external validation patients, ISUP grade group significantly differentiated outcomes. However, among intermediate-risk patients only, with ISUP grade groups 2–3, ISUP itself as well as models M0, M1, and M2 did not differentiate outcomes. In the same cohort, M3 successfully differentiated BCR-related outcomes (*p* < 0.05). The multimodal model M3 was the only model that consistently differentiated outcomes with statistical significance across each cohort and subgroup. Complete clinical-comparison results are displayed in Supplemental Fig. S1. Because post-surgical scores could not be combined across centers, combined pre-surgical ISUP grade-group results are shown in Supplemental Fig. S2. Pre-surgical ISUP grade group significantly differentiated outcomes in the full test-set population (*p* < 0.05), but not within the intermediate-risk subgroup (*p >* 0.05).

### G. Discussion

We present the development and external validation of an automated multimodal deep learning-based model to predict RP treatment outcomes from baseline MRI, age, and PSA. Across both the internal and external validation sets, the fully automated multimodal model M3 was the only model that differentiated those who went on to experience BCR with statistical significance across each center and within intermediate-risk subgroups. Since intermediate-risk patients are the most difficult group in which to predict outcomes based on current gold standards, a model that can reproducibly and accurately predict patient outcomes within this group, independent of current metrics, is a potentially important contribution and could help further delineate patient risk and inform treatment strategies. While reader-based scoring systems such as NCCN groups, Gleason grades, and PI-RADS remain clinically important, their subjective nature introduces variability. An automated approach such as the one presented here could become instrumental, after further multicenter external validation and ethical testing, in informing physicians and patients about individualized surgical-outcome risk.

The field of AI in prostate imaging has attracted substantial attention in the past decade, but existing work often faces barriers to realistic implementation in the clinic. Leading studies frequently rely on reader contours or biopsy grades, and single-center studies lacking external validation remain common. Our radiomics-extraction pipeline is fully automated and compares favorably with CAPRA-S in the internal test cohort and with ISUP grade group in the external validation cohort, providing evidence that these results may generalize and that the extracted features have clinical value. Our findings suggest that AI may further refine prognostic ability independently of existing clinical gold standards, which may be especially helpful for intermediate-risk patients.

Several studies have explored the clinical relevance of quantitative MRI-based assessment, but few take a multimodal approach that integrates clinical covariates [16]. Those that do are generally limited to single-center studies, do not provide evidence that the findings generalize to external centers, or incorporate non-automated features such as contours or biopsy reads that are subject to reader variability [14, 17–19]. Some also focus on comparisons between low/intermediate-versus high-risk groups, rather than the more challenging within-intermediate-risk comparison [16]. More common in the PCa AI literature is a focus on histopathology or integration of pathology and MRI [17, 20]. However, digitized pathology is not necessarily feasible in many clinical settings, and many studies focus on post-operative pathology, which is not relevant for stratifying pre-treatment risk.

Compared with existing literature, few models can be directly compared with ours because our focus is specifically on multimodal, automated, pre-operative MRI-based prognosis in PCa. Our AUC of 0.75 in an external validation set is broadly consistent with the literature while our sample size and external validation provide evidence for generalizability. One similar imaging-only study reported an AUC of 0.73 for predicting BCR [21]. However, its external validation cohort was considerably smaller, with only 50 patients and 7 BCR events. Another study featured an even smaller cohort of only 18 test patients and reported an AUC of 0.73 [22]. Other studies reporting similar or higher AUCs focused solely on single centers or physician reads [14, 23, 24], highlighting continued concerns about reproducibility and bias [18, 24]. The fact that M3 can differentiate intermediate-risk outcomes with significance in an external center in a completely automated fashion, and thus without reader variability, is therefore encouraging. Still, multicenter analyses will be imperative to ensure fair and comprehensive efficacy.

Although the main focus of this study is not explainability, the weighted feature importances offer insight into the features the algorithm relies on for decision-making. PSA was the most heavily weighted feature in models M0, M1, and M3. In addition, radiomics features associated with heterogeneity, disorder, and tumor size and shape were heavily weighted in models M2 and M3. A more detailed discussion of these features is provided in the supplementary discussion.

Although the results indicate that M3 may provide value in PCa prognostication beyond existing metrics, several aspects of the study limit its application and call for further work before clinical implementation. First, information was not available to calculate CAPRA-S in center 2, nor was information available regarding adjuvant radiotherapy. Second, the model itself did not account for time to BCR, although the BCR-free survival analysis did. Third, although external validation provides some evidence of generalizability, the study relies on only two centers, and variability across clinical environments remains an open question. Larger multicenter or national studies will be needed to verify that these results remain clinically reliable regardless of center demographics and imaging technology.

## IV. CONCLUSIONS

We present a fully automated deep learning-based multimodal model that predicts biochemical recurrence after radical prostatectomy in prostate cancer patients using baseline imaging and routinely available clinical covariates. The model demonstrated promising generalizability in an external validation center and was the only evaluated model that consistently differentiated outcomes in intermediate-risk patients across both centers. These findings support continued multicenter validation of imaging-based multimodal AI for individualized treatment-outcome prediction in prostate cancer.

## Data Availability

All data (results but not patient data) produced in the present study are available upon reasonable request to the authors.

https://github.com/NIH-MIP/MultimodalRadiomicsSurgeryModel

## A. SUPPLEMENTARY MATERIALS

### 1. Supplemental Methods

#### a. Development Cohort: Center 1 Image Acquisition

MRIs were obtained at 3 T (Achieve 3.0 T TX, Philips Healthcare) using a 16-channel phase-array surface coil with or without endorectal coils (BPX-30, Medrad). An expert genitourinary radiologist with 20 years of experience evaluated all scans prospectively from 2008 to 2018. Scans obtained between May 2015 and 2018 were prospectively evaluated according to PI-RADS v2.0, and those obtained between January 2008 and May 2015 were retrospectively re-evaluated using PI-RADS v2.0. Imaging sequences included T2-weighted imaging, apparent diffusion coefficient (ADC) maps, high-*b*-value diffusion-weighted imaging (*b* = 2000 or 1500 s/mm^2^), and dynamic contrast-enhanced MRI. For some patients (prior to December 2011), MRI acquisition did not include high-*b*-value DWI, so these were calculated at *b* = 1500 s/mm^2^. DCE MRI was not used in the present analysis.

#### b. Center 1 Biopsy Readers and Methods

An expert urologist with 23 years of experience and an expert interventional radiologist with 21 years of experience performed all biopsies transrectally or transperineally. Surgical procedures were carried out robotically (Da Vinci) by the same urologist and another urologist with 6 years of experience. An expert genitourinary pathologist with 45 years of experience evaluated all biopsy and RP specimens while blinded to MRI reports.

#### c. External Validation Cohort: Center 2 Image Acquisition

MRIs at center 2 were obtained at either 1.5 T (Magnetom Aera, Siemens Healthcare; Achieva, Philips Healthcare; or Signa Explorer 1.5T, GE Healthcare; *n* = 188) or 3 T (Signa Architect 3T, GE Healthcare; *n* = 110). PI-RADS v2.1 scoring was prospectively assessed by two radiologists with more than 10 years of experience reading prostate MRI. See Fig. 1 for the patient-population criteria at each center.

#### d. Center 2 Biopsy Readers and Methods

An expert urologist with 15 years of experience and three expert interventional radiologists, each with at least 10 years of experience, performed all biopsies transrectally or transperineally. Sixty percent of cases were performed robotically (Da Vinci Xi), and 40% were done using a conventional approach by the same urologist. An expert genitourinary pathologist with 10 years of experience evaluated all biopsy and RP specimens.

### 1. Supplementary Discussion: Feature Importance

Inspection of feature importances across models shows that PSA is unsurprisingly, but still notably, the most heavily weighted feature in models M0, M1, and M3. Radiomics features across M2 and M3 demonstrate some overlapping prioritizations. M2 heavily weights GLSZM Zone Variance, which captures variability in the size of homogeneous zones within the lesion. Additional texture features include GLSZM Small Area Emphasis and GLRLM Run Length Non-Uniformity, both of which relate to textural irregularity and non-uniformity of the lesion. M3 included GLSZM Gray Level Variance, quantifying variability in gray-level intensities across homogeneous zones, potentially indicating that the model is capturing differences in tumor heterogeneity. Other features in M2, such as Shape Sphericity and First Order Range, and in M3, such as Least Axis Length, Maximum 2D Diameter, and Mesh Volume, plausibly relate to tumor size, bulk, and heterogeneity. Although the exact features differ between models, a core set of radiomics domains—texture, shape, and intensity—appears to encode biologically meaningful signals that a radiologist would also recognize. This convergence supports the clinical relevance of these radiomics features for individualized risk assessment in PCa.

**Table S1. Supplemental Table S1.**
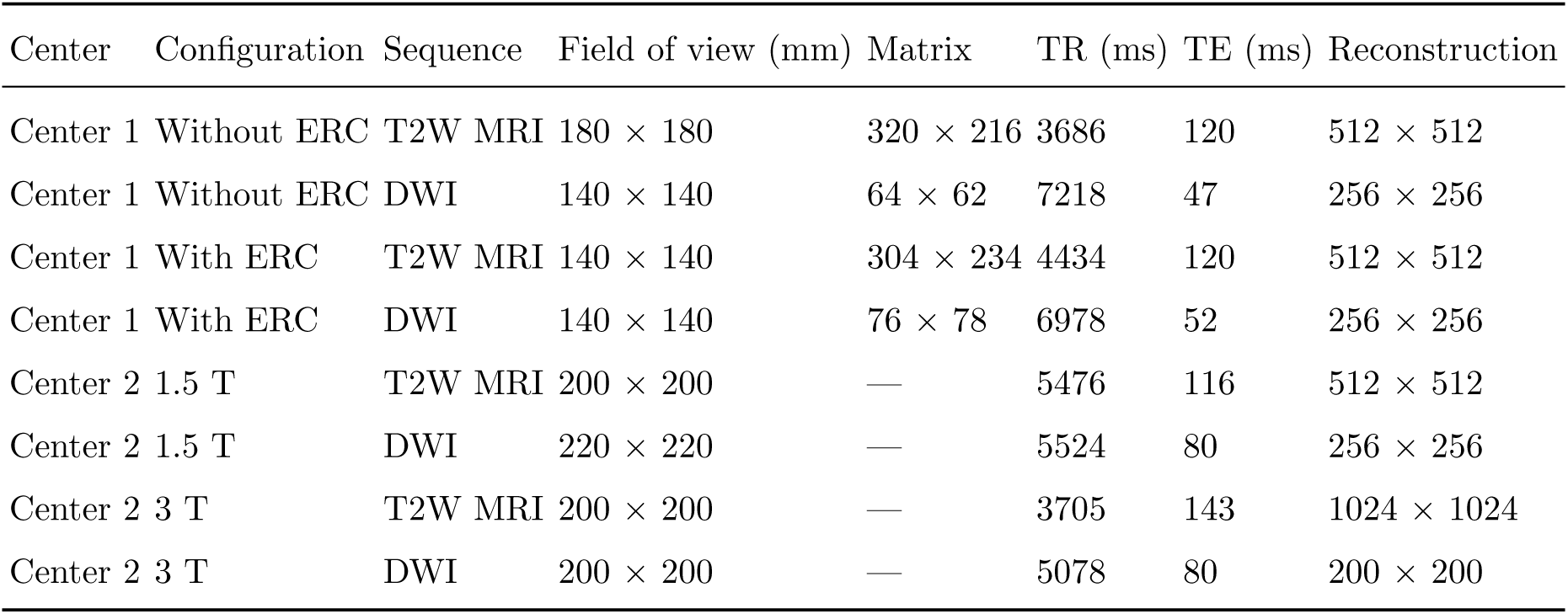
Imaging parameters describing biparametric MRI specifications across each center.

| Center | Configuration | Sequence | Field of view (mm) | Matrix | TR (ms) | TE (ms) | Reconstruction |
| --- | --- | --- | --- | --- | --- | --- | --- |
| Center 1 | Without ERC | T2W MRI | $180 \times 180$ | $320 \times 216$ | 3686 | 120 | $512 \times 512$ |
| Center 1 | Without ERC | DWI | $140 \times 140$ | $64 \times 62$ | 7218 | 47 | $256 \times 256$ |
| Center 1 | With ERC | T2W MRI | $140 \times 140$ | $304 \times 234$ | 4434 | 120 | $512 \times 512$ |
| Center 1 | With ERC | DWI | $140 \times 140$ | $76 \times 78$ | 6978 | 52 | $256 \times 256$ |
| Center 2 | 1.5 T | T2W MRI | $200 \times 200$ | — | 5476 | 116 | $512 \times 512$ |
| Center 2 | 1.5 T | DWI | $220 \times 220$ | — | 5524 | 80 | $256 \times 256$ |
| Center 2 | 3 T | T2W MRI | $200 \times 200$ | — | 3705 | 143 | $1024 \times 1024$ |
| Center 2 | 3 T | DWI | $200 \times 200$ | — | 5078 | 80 | $200 \times 200$ |

**Figure S1.**
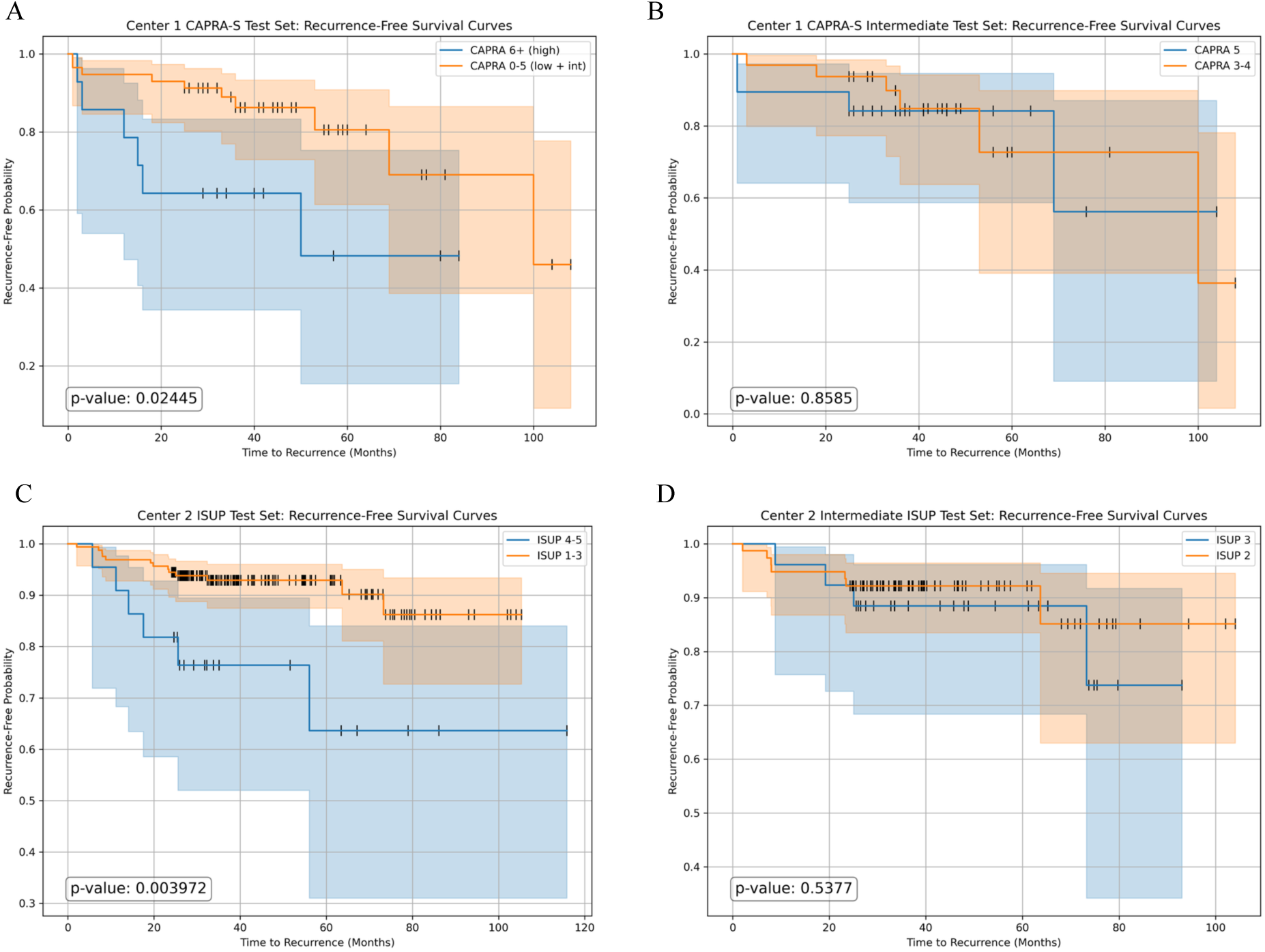
Clinical comparison by center. Recurrence-free survival curves with associated log-rank tests for clinical baselines in each cohort.

**Figure S2.**
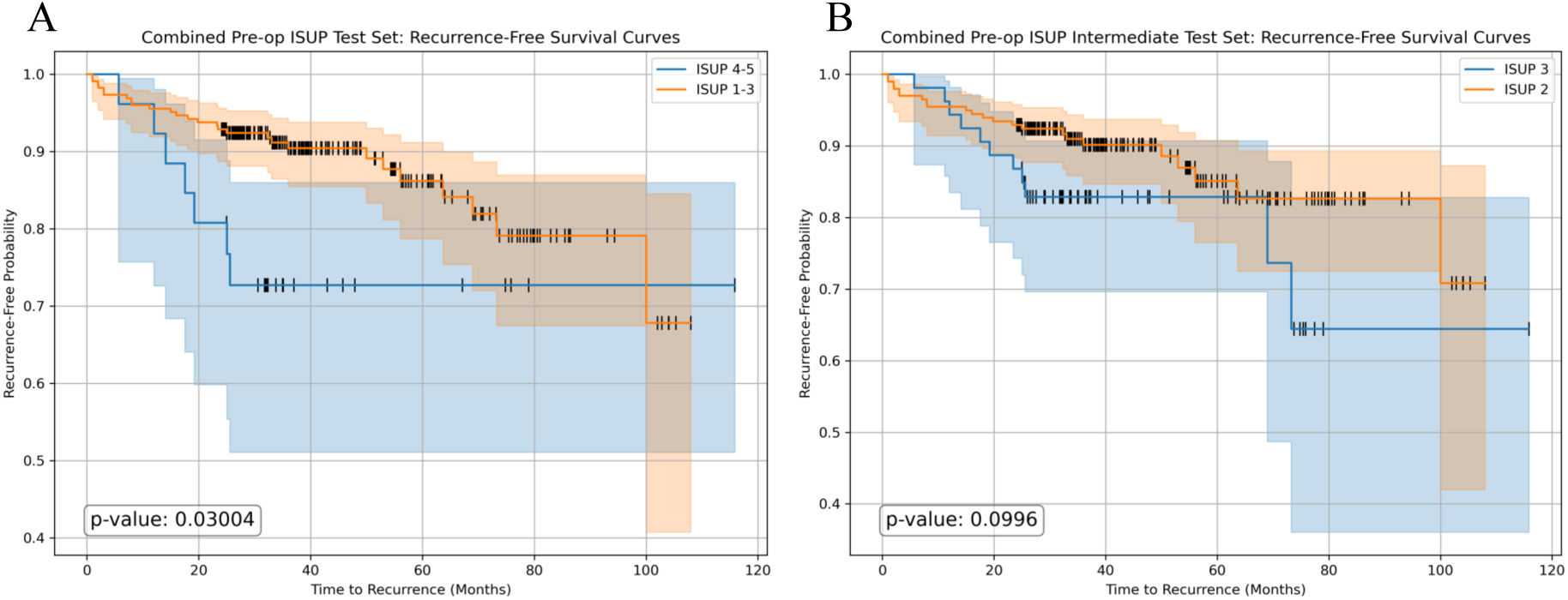
Pre-surgical clinical comparison. Recurrence-free survival curves with associated log-rank tests for pre-surgical biopsy ISUP grade-group comparisons in the combined test-set cohort.

